# Reliable prediction of childhood obesity using only routinely collected EHRs is possible

**DOI:** 10.1101/2024.01.29.24301945

**Authors:** Mehak Gupta, Thao-Ly T. Phan, Daniel Eckrich, H. Timothy Bunnell, Rahmatollah Beheshti

## Abstract

**Objective:** Identifying children at high risk of developing obesity can offer a critical time to change the course of the disease before it establishes. Numerous studies have tried to achieve this; but practical limitations remain, including (i) relying on data not present in routinely available pediatric data (like prenatal data), (ii) focusing on a single age prediction (hence, not tested across ages), and (iii) not achieving good results or adequately validating those.

**Methods:** A customized sequential deep learning model was built to predict the risk of childhood obesity, focusing especially on capturing the temporal patterns. The model was trained only on routinely collected EHRs, containing a list of features identified by a group of clinical experts, and sourced from 36,191 diverse children aged 0 to 10. The model was evaluated using extensive discrimination, calibration, and utility analysis; and was validated temporally, geographically, and across various subgroups.

**Results:** Our results are mostly better (and never worse) than all previous studies, including those that focus on single-age predictions or link EHRs to external data. Specifically, the model consistently achieved an area under the curve (AUROC) of above 0.8 (with most cases around 0.9) for predicting obesity within the next 3 years for children 2 to 7. The validation results show the robustness of the model. Furthermore, the most influential predictors of the model match important risk factors of obesity.

**Conclusions:** Our model is able to predict the risk of obesity for young children using only routinely collected EHR data, greatly facilitating its integration with the periodicity schedule. The model can serve as an objective screening tool to inform prevention efforts, especially by helping with very delicate interactions between providers and families in primary care settings.

## 1. Introduction

Childhood obesity is a major public health problem across the globe and in the US. Obesity affects almost 1 in 5 and about 14.7 million children and adolescents, for children and adolescents of 2-19 years of age [1]. Despite evidence of potentially modifiable risk factors that contribute to the development of obesity in children, children are often referred for obesity interventions after their obesity is well established and when intervention is less likely to be successful [2]. Prior studies show that less than 30% of children with overweight or obesity are identified by their provider at primary care visits and less than 10% of children have a diagnoses code for overweight or obesity placed during visits [3, 4, 5, 6]. While pediatric providers frequently use recommended CDC or WHO age and sex-specific BMI charts, they do not often recognize or address weight concerns until children cross overweight and obesity thresholds on these charts [7]. Primary barriers to effective diagnoses and management of obesity in pediatric systems are reported as lack of time, limited resources, and uncertainties about the level of risk [6, 8].

In such a context, reliable predictive models of childhood obesity integrated within the structure of the common well-child visits have the potential to provide timely risk alerts and inform more effective interventions to prevent and control this disease. Clinical predictive models for obesity, designed using artificial intelligence and machine learning (AI/ML) methods, are being considered to understand the contributing factors to the obesity epidemic and inform more effective interventions [9, 10, 11, 12, 13, 14, 15, 16, 17, 18].

One major limitation of existing obesity prediction models is that they use features that, despite their importance, are generally not available in EHRs or are difficult and time-consuming to collect, such as genetic background, parental data, and children’s habits. Another limitation is that they mostly focus on predicting the risk of obesity at only one age point (e.g., at age five) and are not flexible in allowing obesity screening for patients at different age points. Additionally, large-scale and rigorously validated predictive models of obesity are rare.

In this work, we aim to demonstrate the possibility of achieving reliable estimates of the future risks of childhood obesity using commonly available (unaugmented) EHR elements across multiple age ranges. As children’s weight status in early childhood is highly predictive of weight status into adulthood [19], we focus our study on children up until the age of 10 years. This age range also includes a few years after the “adiposity rebound” patterns observed in children’s weight [20, 21]. We rigorously evaluate our model through an extensive series of temporal, geographic, and subgroup validations and explore the most important predictors of obesity in our model. The overarching aim of our study is to offer a general (practice-agnostic) predictive model that can be integrated within any healthcare system’s EHR to reliably predict a child’s risk for obesity throughout early childhood to support clinical decision-making and obesity prevention at the point of care.

## 2. Materials and Methods

### 2.1 Data source and study cohort selection

The EHR data used in this analysis was extracted from Nemours Children’s Health; a large pediatric healthcare network in the United States (US) primarily spanning the states of Delaware, Florida, Maryland, New Jersey, and Pennsylvania. The IRB panel at Nemours approved this research study. We used encounters between January 1, 2002, to December 31, 2019, obtained prior to the pandemic to avoid any short-term influence that the pandemic had on weight gain patterns[22]. A total of 68 029 children with 44 401 791 encounters were selected as having: (i) no evidence of type 1 diabetes, and (ii) no evidence of cancer, and sickle cell disease. Of them, 37 844 children were included having at least two routine infant checkups and at least one checkup between 2 to age 10 with recorded weight and length/height measurements. Of them, 1 653 children were excluded whose year of birth could not be verified, leaving 36 191 children. The final cohort was separately divided for temporal and geographic validation. More details about the Nemours EHR dataset are provided in Supplementary A. Figure 1 shows the steps we took to extract our cohort of 36 191 patients for the model construction.

**Figure 1:**
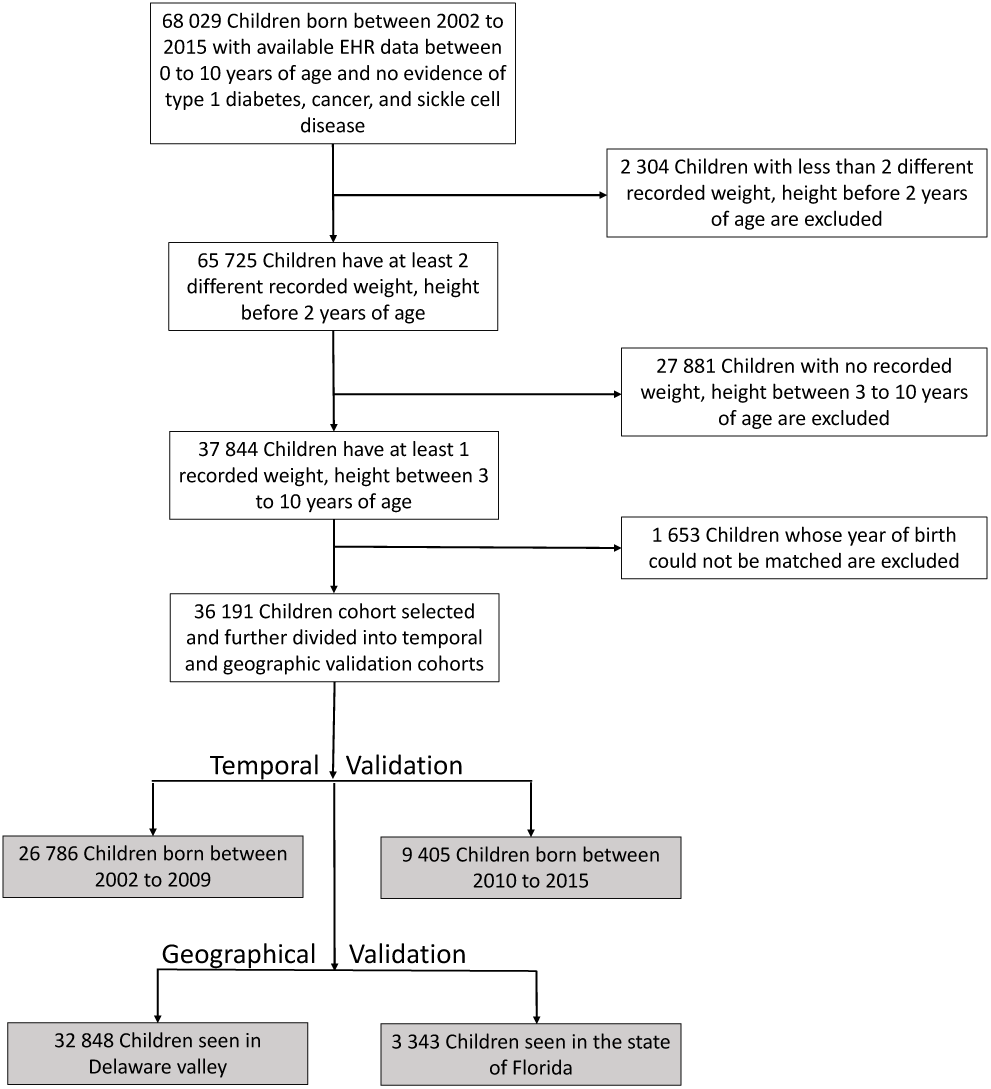
The cohort selection steps. Temporal and geographic validation regimes are shown in gray.

### 2.2 Feature selection and data preprocessing

We extracted clinically-relevant features to childhood obesity, by using a data-driven approach coupled with input from clinical experts [23]. Details about this process are presented in Supplementary B. The final EHR features consisted of 138 diagnoses (patient conditions and family-history conditions associated with obesity), 84 ATC3 (Anatomical Therapeu-tic Chemical Classification Level-3) medication groups, and 51 measurements (vitals and labs).

Following prior studies [24], the EHR features before age 2, were segmented into 5 windows. These 5 sliding windows correspond to 0–4 months, 4–8 months, 8–12 months, 12–18 months, and 18–24 months. All EHR features after age 2 were segmented into 1-year windows due to the lower frequency of medical visits (compared to before 2).

Weight and height data were converted to Weight-for-length (WFL) and BMI% as defined by WHO and CDC, respectively. these values were considered Missing Completely at Random (MCAR) and imputed with carry-forward of the most recent value. WFL and BMI% were categorized into underweight, normal, overweight, and obesity categories. We defined cutoffs for normal weight, overweight, and obesity in accordance with the CDC’s standard thresholds of the 85^th^ and 95^th^ percentiles for overweight and obesity, respectively. Because WFL% and BMI% trajectories can be helpful for determining the risk of obesity [25], we also engineered a WFL% change feature that calculates the change in WFL% values between 5 windows defined above for model input. Table 1 shows the characteristics of the final cohort used.

**Table 1:**
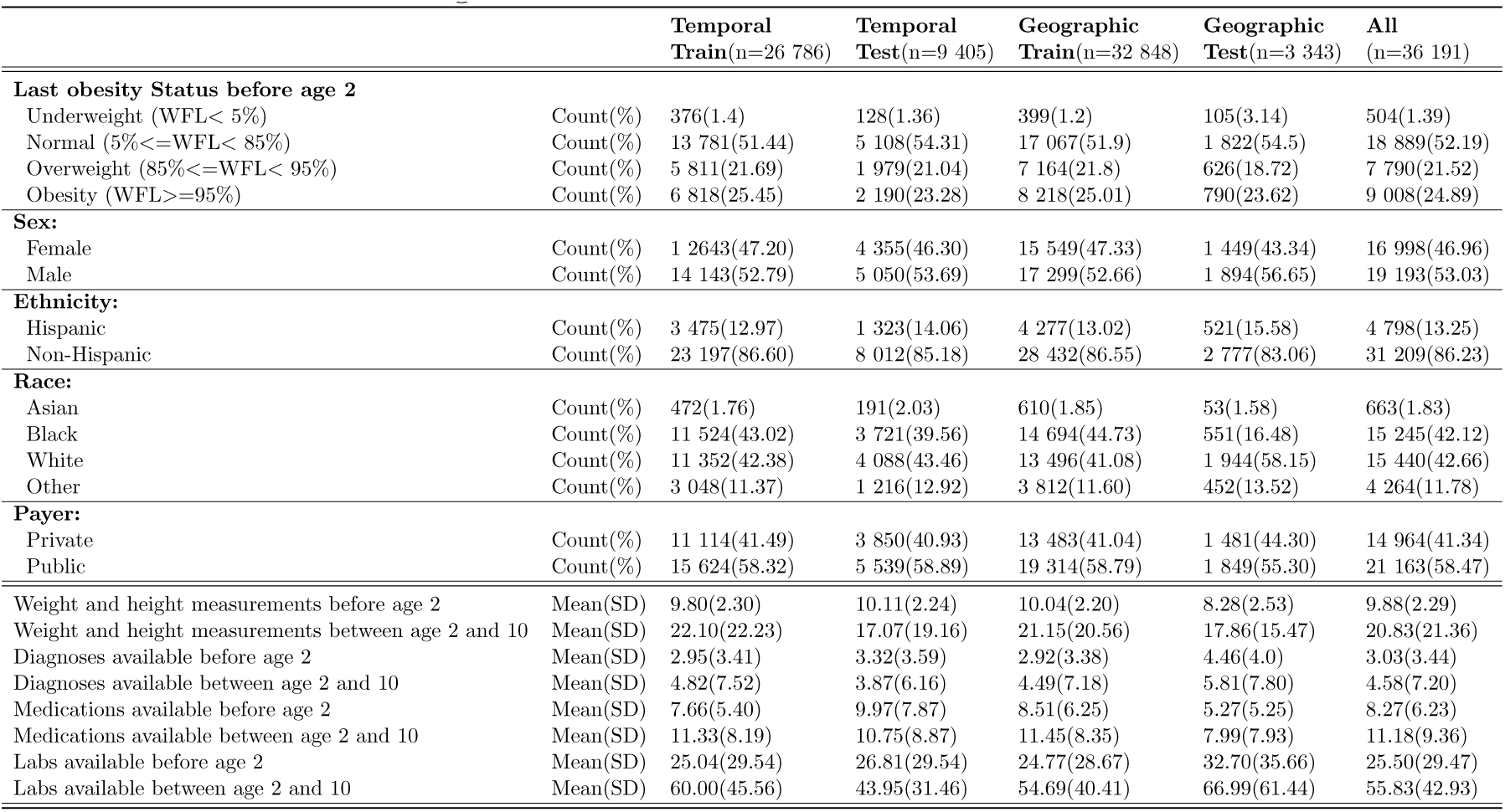
Statistics for the study cohort (full), as well as the cohorts used for temporal and geographic validation. Counts with the percentage prevalence in the respective cohort for last obesity status before age 2, sex, race, ethnicity, and payer are shown. In the bottom part, the mean(standard deviation) for the number of times that weight, height, diagnoses, medications, and lab measurements were recorded before and after age 2 are shown.

|  |  | Temporal<br>Train(n=26 786) | Temporal<br>Test(n=9 405) | Geographic<br>Train(n=32 848) | Geographic<br>Test(n=3 343) | All<br>(n=36 191) |
| --- | --- | --- | --- | --- | --- | --- |
| **Last obesity Status before age 2** |  |  |  |  |  |  |
| Underweight (WFL< 5%) | Count(%) | 376(1.4) | 128(1.36) | 399(1.2) | 105(3.14) | 504(1.39) |
| Normal (5%<=WFL< 85%) | Count(%) | 13 781(51.44) | 5 108(54.31) | 17 067(51.9) | 1 822(54.5) | 18 889(52.19) |
| Overweight (85%<=WFL< 95%) | Count(%) | 5 811(21.69) | 1 979(21.04) | 7 164(21.8) | 626(18.72) | 7 790(21.52) |
| Obesity (WFL>=95%) | Count(%) | 6 818(25.45) | 2 190(23.28) | 8 218(25.01) | 790(23.62) | 9 008(24.89) |
| **Sex:** |  |  |  |  |  |  |
| Female | Count(%) | 1 2643(47.20) | 4 355(46.30) | 15 549(47.33) | 1 449(43.34) | 16 998(46.96) |
| Male | Count(%) | 14 143(52.79) | 5 050(53.69) | 17 299(52.66) | 1 894(56.65) | 19 193(53.03) |
| **Ethnicity:** |  |  |  |  |  |  |
| Hispanic | Count(%) | 3 475(12.97) | 1 323(14.06) | 4 277(13.02) | 521(15.58) | 4 798(13.25) |
| Non-Hispanic | Count(%) | 23 197(86.60) | 8 012(85.18) | 28 432(86.55) | 2 777(83.06) | 31 209(86.23) |
| **Race:** |  |  |  |  |  |  |
| Asian | Count(%) | 472(1.76) | 191(2.03) | 610(1.85) | 53(1.58) | 663(1.83) |
| Black | Count(%) | 11 524(43.02) | 3 721(39.56) | 14 694(44.73) | 551(16.48) | 15 245(42.12) |
| White | Count(%) | 11 352(42.38) | 4 088(43.46) | 13 496(41.08) | 1 944(58.15) | 15 440(42.66) |
| Other | Count(%) | 3 048(11.37) | 1 216(12.92) | 3 812(11.60) | 452(13.52) | 4 264(11.78) |
| **Payer:** |  |  |  |  |  |  |
| Private | Count(%) | 11 114(41.49) | 3 850(40.93) | 13 483(41.04) | 1 481(44.30) | 14 964(41.34) |
| Public | Count(%) | 15 624(58.32) | 5 539(58.89) | 19 314(58.79) | 1 849(55.30) | 21 163(58.47) |
| Weight and height measurements before age 2 | Mean(SD) | 9.80(2.30) | 10.11(2.24) | 10.04(2.20) | 8.28(2.53) | 9.88(2.29) |
| Weight and height measurements between age 2 and 10 | Mean(SD) | 22.10(22.23) | 17.07(19.16) | 21.15(20.56) | 17.86(15.47) | 20.83(21.36) |
| Diagnoses available before age 2 | Mean(SD) | 2.95(3.41) | 3.32(3.59) | 2.92(3.38) | 4.46(4.0) | 3.03(3.44) |
| Diagnoses available between age 2 and 10 | Mean(SD) | 4.82(7.52) | 3.87(6.16) | 4.49(7.18) | 5.81(7.80) | 4.58(7.20) |
| Medications available before age 2 | Mean(SD) | 7.66(5.40) | 9.97(7.87) | 8.51(6.25) | 5.27(5.25) | 8.27(6.23) |
| Medications available between age 2 and 10 | Mean(SD) | 11.33(8.19) | 10.75(8.87) | 11.45(8.35) | 7.99(7.93) | 11.18(9.36) |
| Labs available before age 2 | Mean(SD) | 25.04(29.54) | 26.81(29.54) | 24.77(28.67) | 32.70(35.66) | 25.50(29.47) |
| Labs available between age 2 and 10 | Mean(SD) | 60.00(45.56) | 43.95(31.46) | 54.69(40.41) | 66.99(61.44) | 55.83(42.93) |

The EHR data also included demographic information about sex (male or female), race (categorized as White, Black, Asian, and other), ethnicity (categorized as Hispanic or non-Hispanic), and payer (categorized as public and private insurance). We also included the Child Opportunity Index (COI) by geolocating the last address of patients before age 2 [26]. COI combines indicators of educational opportunity, health and environment, and socioeconomic opportunity for all US neighborhoods. COI ranges from 1-100, with higher numbers representing neighborhoods with more opportunities. Details about demographic feature generation are provided in Supplementary B.1.

A binary representation (not MCAR) was used for all features, where the presence of a value for the variable was captured with a 1, and 0 otherwise. We used quintile binning for all numeric measurement features to divide each measurement into 5 categorical features. In total, we generated 506 binary features: 138 (diagnoses) + 84 (medications) + 51*5 (measurements in 5 percentile bins) + 10 (demographic categories) + 4 (underweight, normal, overweight, and obesity) + 5 (WFL% changes in 5 percentile bins) + 10 (COIs in 10 percentile bins). Please refer to Table S1 in Supplementary B.5 for a detailed list of the final features.

### 2.3 Descriptive analysis

To study the transition of obesity status, we analyzed the distribution of children with BMI% *≥* 95 at age 3 to 10 that transitioned from different WFL% status (WFL% *<* 5, 5 *≤* WFL% *<* 85, 85 *≤* WFL% *<* 95, and WFL% *≥* 95) at age 2. Figure 2 shows that 22% of children with obesity at age 3 had normal weight at infancy and this percentage increases with increasing age, where nearly half (49%) of children with obesity at age 10 had normal weight at infancy.

**Figure 2:**
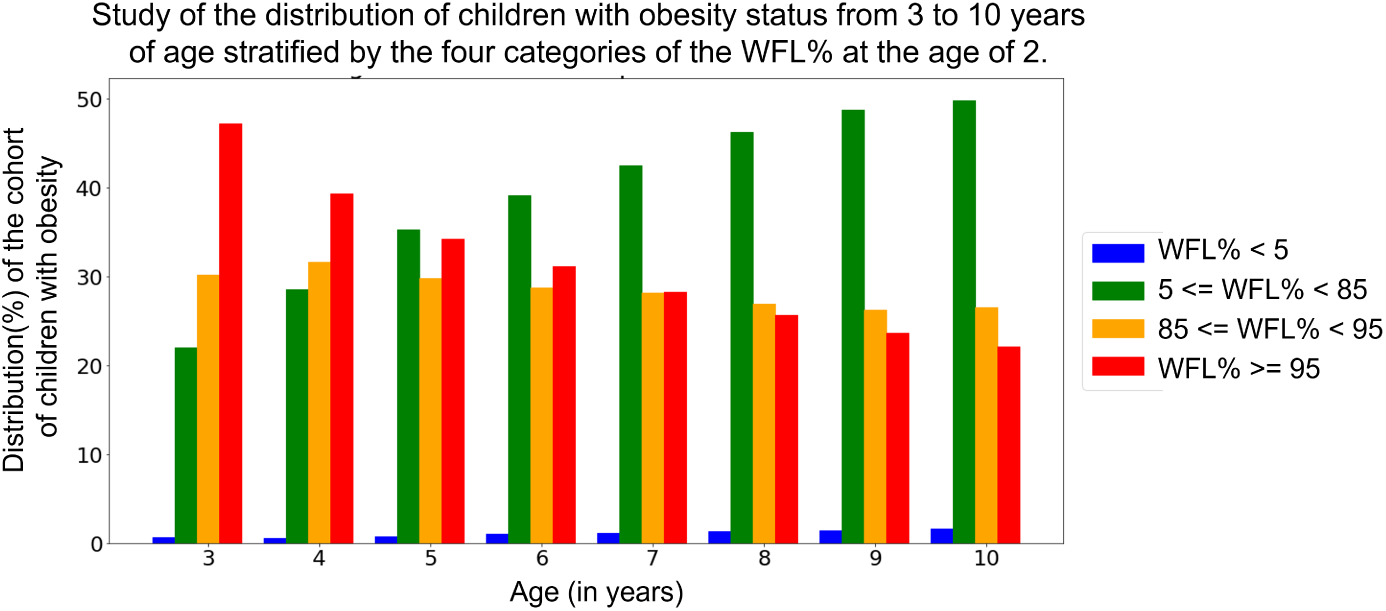
Study of the distribution of children with obesity (BMI% *≥* 95) at age 3 to 10. We analyze what percentage of children with obesity at ages 3 to 10 had WFL% *<* 5, 5 *≤* WFL% *<* 85, 85 *≤* WFL% *<* 95, and WFL% *≥* 95 at age 2. X-axis shows the distribution of the cohort with obesity at every age from 3 to age 10. Four categories of WFL% at age 2 are shown on the right.

### 2.4 Deep learning model training

We adopted our encoder-decoder^1^ deep neural network model and the training procedure presented in detail in prior work [27, 18]. The encoder part consists of long short-term memory (LSTM)^2^ cells and the decoder consists of a feed-forward network with two fully-connected layers.

To train the model, we first trained the encoder network on the input EHR data from 0 to 2 years. EHR data for every year after age 2 until the age that the prediction was performed at (i.e., the end of the observation window) were then combined (concatenated) with the *n*-dimensional vector representation derived from the encoder for 0-2 years of EHR data. The number of years of data after age 2 depends on the length of the patient’s recorded medical history. Using this design, all the prior data for each patient was used for model training to predict the risk of obesity for the next 3 years. A detailed description of the architecture design and parameter settings is provided in Supplementary C.

## 3. Experiments and Results

### 3.1 Setup

We extensively evaluated our predictive model by studying its discrimination power, calibration, and robustness. We validated our model temporally and geographically and studied the model’s performance across subpopulations (which can also capture the model’s fairness across these groups), using separate test datasets. When using the entire cohort, the data was split with an 80:20 train and test regime, with 5% of the training data as a validation set to fix the best model. Model performance was reported exclusively on the test dataset. The confidence intervals (CI) are calculated using 100 bootstrapped replicates.

#### Baseline comparison

Similar to prior work [28, 11], we consider only using the last WFL% (below 2 years) and BMI% (above 2 years) available in the observation window as a baseline. This scenario mimics what is generally used in clinical practice for screening children (i.e., only using present data).

#### Discrimination power

We report prediction performance using Area Under Receiver Operating Characteristic (AUROC), Area Under Precision-Recall Curve (AUPRC), sensitivity, and specificity. We provide sensitivity and specificity at different binary classification thresholds, demonstrating the trade-off between false negatives (patients who develop obesity but are not predicted to develop obesity by the model) and false positives (patients who do not develop obesity but are predicted to develop obesity by the model). Specifically, we fixed either sensitivity or specificity at 90% and 95% and measured the value of the other metric.

#### Utility and calibration

We perform decision curve analysis [29] to demonstrate the tradeoffs (costs and benefits) of using the prediction model to analyze the clinical utility at various thresholds. Related to the above approach, we provide calibration metrics to help quantify how well the predicted probabilities of an outcome match the true probabilities observed in the data [30, 31].

#### Temporal validation

To study our model’s robustness across time shifts, we divided our cohort according to the date of birth of the children. The data for 26 786 children who were born between January 1, 2002, and December 31 2009, were included as a training set; and the data for 9 405 children who were born between January 1, 2010, and December 31, 2015, were included in the test set. We report AUROC for the temporal validation in Table 2.

**Table 2:**
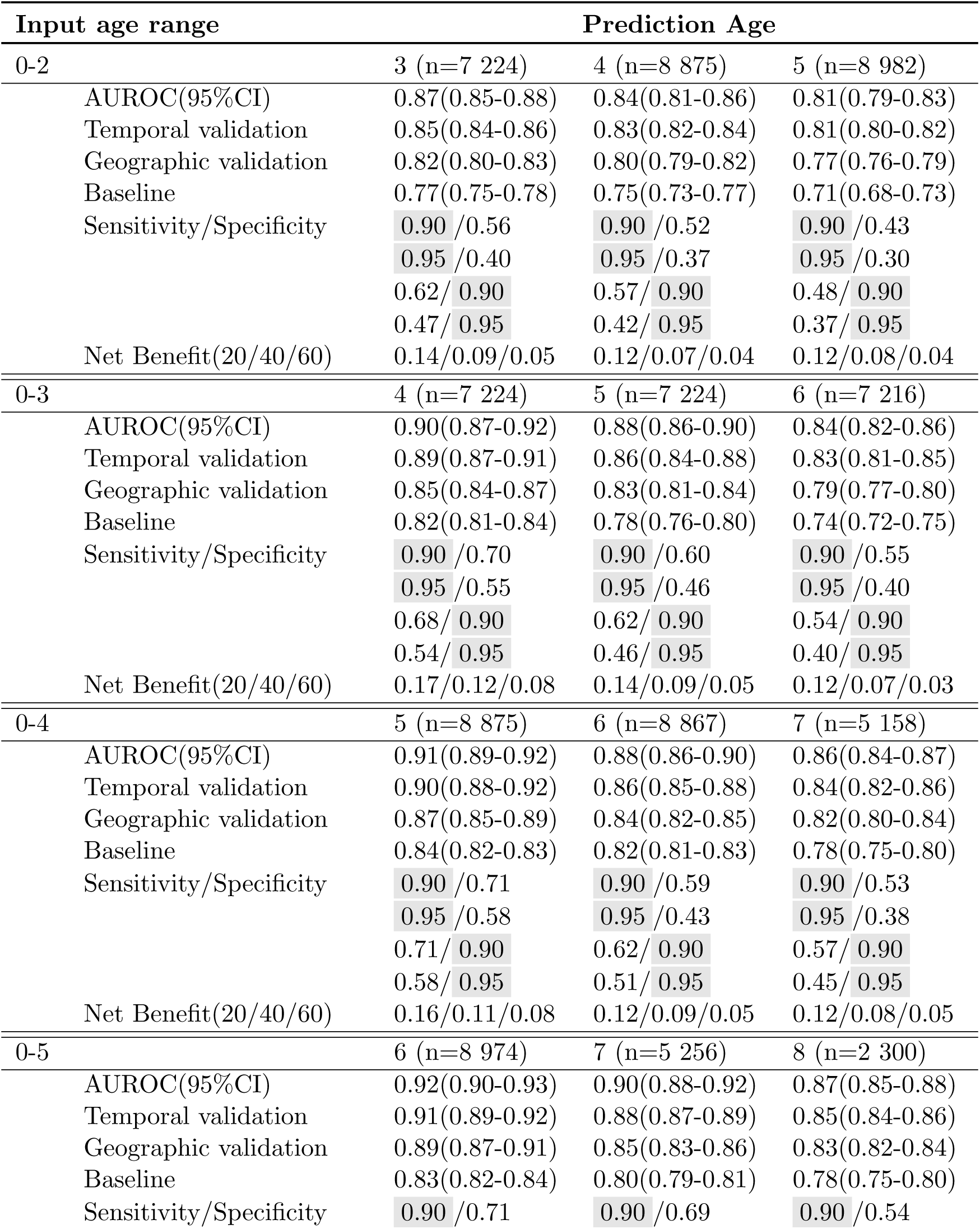
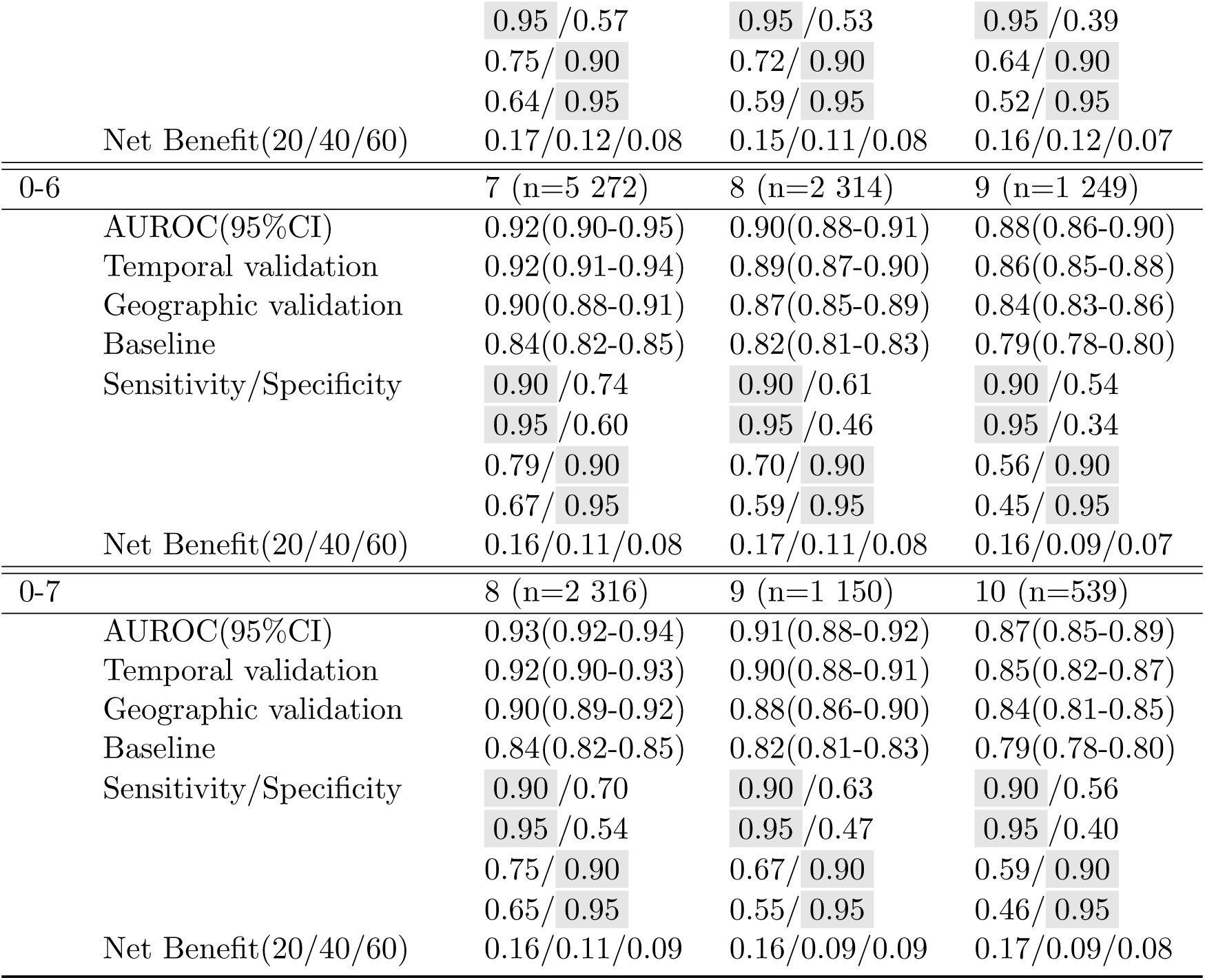
Predictive performance across six different observation windows. Results are shown for prediction for the next 1, 2, and 3 years. The fixed values for sensitivity and specificity are highlighted. The net benefits are shown at three thresholds of 20, 40, and 60 percent.

| Input age range | Prediction Age |  |  |
| --- | --- | --- | --- |
| 0-2 | 3 (n=7 224) | 4 (n=8 875) | 5 (n=8 982) |
| AUROC(95%CI) | 0.87(0.85-0.88) | 0.84(0.81-0.86) | 0.81(0.79-0.83) |
| Temporal validation | 0.85(0.84-0.86) | 0.83(0.82-0.84) | 0.81(0.80-0.82) |
| Geographic validation | 0.82(0.80-0.83) | 0.80(0.79-0.82) | 0.77(0.76-0.79) |
| Baseline | 0.77(0.75-0.78) | 0.75(0.73-0.77) | 0.71(0.68-0.73) |
| Sensitivity/Specificity | 0.90 /0.56<br>0.95 /0.40 | 0.90 /0.52<br>0.95 /0.37 | 0.90 /0.43<br>0.95 /0.30 |
|  | 0.62/ 0.90 | 0.57/ 0.90 | 0.48/ 0.90 |
|  | 0.47/ 0.95 | 0.42/ 0.95 | 0.37/ 0.95 |
| Net Benefit(20/40/60) | 0.14/0.09/0.05 | 0.12/0.07/0.04 | 0.12/0.08/0.04 |
| 0-3 | 4 (n=7 224) | 5 (n=7 224) | 6 (n=7 216) |
| AUROC(95%CI) | 0.90(0.87-0.92) | 0.88(0.86-0.90) | 0.84(0.82-0.86) |
| Temporal validation | 0.89(0.87-0.91) | 0.86(0.84-0.88) | 0.83(0.81-0.85) |
| Geographic validation | 0.85(0.84-0.87) | 0.83(0.81-0.84) | 0.79(0.77-0.80) |
| Baseline | 0.82(0.81-0.84) | 0.78(0.76-0.80) | 0.74(0.72-0.75) |
| Sensitivity/Specificity | 0.90 /0.70<br>0.95 /0.55 | 0.90 /0.60<br>0.95 /0.46 | 0.90 /0.55<br>0.95 /0.40 |
|  | 0.68/ 0.90 | 0.62/ 0.90 | 0.54/ 0.90 |
|  | 0.54/ 0.95 | 0.46/ 0.95 | 0.40/ 0.95 |
| Net Benefit(20/40/60) | 0.17/0.12/0.08 | 0.14/0.09/0.05 | 0.12/0.07/0.03 |
| 0-4 | 5 (n=8 875) | 6 (n=8 867) | 7 (n=5 158) |
| AUROC(95%CI) | 0.91(0.89-0.92) | 0.88(0.86-0.90) | 0.86(0.84-0.87) |
| Temporal validation | 0.90(0.88-0.92) | 0.86(0.85-0.88) | 0.84(0.82-0.86) |
| Geographic validation | 0.87(0.85-0.89) | 0.84(0.82-0.85) | 0.82(0.80-0.84) |
| Baseline | 0.84(0.82-0.83) | 0.82(0.81-0.83) | 0.78(0.75-0.80) |
| Sensitivity/Specificity | 0.90 /0.71<br>0.95 /0.58 | 0.90 /0.59<br>0.95 /0.43 | 0.90 /0.53<br>0.95 /0.38 |
|  | 0.71/ 0.90 | 0.62/ 0.90 | 0.57/ 0.90 |
|  | 0.58/ 0.95 | 0.51/ 0.95 | 0.45/ 0.95 |
| Net Benefit(20/40/60) | 0.16/0.11/0.08 | 0.12/0.09/0.05 | 0.12/0.08/0.05 |
| 0-5 | 6 (n=8 974) | 7 (n=5 256) | 8 (n=2 300) |
| AUROC(95%CI) | 0.92(0.90-0.93) | 0.90(0.88-0.92) | 0.87(0.85-0.88) |
| Temporal validation | 0.91(0.89-0.92) | 0.88(0.87-0.89) | 0.85(0.84-0.86) |
| Geographic validation | 0.89(0.87-0.91) | 0.85(0.83-0.86) | 0.83(0.82-0.84) |
| Baseline | 0.83(0.82-0.84) | 0.80(0.79-0.81) | 0.78(0.75-0.80) |
| Sensitivity/Specificity | 0.90 /0.71 | 0.90 /0.69 | 0.90 /0.54 |

OBESITY PREDICTION
|  |  |  |  |  |
| --- | --- | --- | --- | --- |
|  |  | 0.95 /0.57 | 0.95 /0.53 | 0.95 /0.39 |
|  |  | 0.75/ 0.90 | 0.72/ 0.90 | 0.64/ 0.90 |
|  |  | 0.64/ 0.95 | 0.59/ 0.95 | 0.52/ 0.95 |
|  | Net Benefit(20/40/60) | 0.17/0.12/0.08 | 0.15/0.11/0.08 | 0.16/0.12/0.07 |
| 0-6 |  | 7 (n=5 272) | 8 (n=2 314) | 9 (n=1 249) |
|  | AUROC(95%CI) | 0.92(0.90-0.95) | 0.90(0.88-0.91) | 0.88(0.86-0.90) |
|  | Temporal validation | 0.92(0.91-0.94) | 0.89(0.87-0.90) | 0.86(0.85-0.88) |
|  | Geographic validation | 0.90(0.88-0.91) | 0.87(0.85-0.89) | 0.84(0.83-0.86) |
|  | Baseline | 0.84(0.82-0.85) | 0.82(0.81-0.83) | 0.79(0.78-0.80) |
|  | Sensitivity/Specificity | 0.90 /0.74 | 0.90 /0.61 | 0.90 /0.54 |
|  |  | 0.95 /0.60 | 0.95 /0.46 | 0.95 /0.34 |
|  |  | 0.79/ 0.90 | 0.70/ 0.90 | 0.56/ 0.90 |
|  |  | 0.67/ 0.95 | 0.59/ 0.95 | 0.45/ 0.95 |
|  | Net Benefit(20/40/60) | 0.16/0.11/0.08 | 0.17/0.11/0.08 | 0.16/0.09/0.07 |
| 0-7 |  | 8 (n=2 316) | 9 (n=1 150) | 10 (n=539) |
|  | AUROC(95%CI) | 0.93(0.92-0.94) | 0.91(0.88-0.92) | 0.87(0.85-0.89) |
|  | Temporal validation | 0.92(0.90-0.93) | 0.90(0.88-0.91) | 0.85(0.82-0.87) |
|  | Geographic validation | 0.90(0.89-0.92) | 0.88(0.86-0.90) | 0.84(0.81-0.85) |
|  | Baseline | 0.84(0.82-0.85) | 0.82(0.81-0.83) | 0.79(0.78-0.80) |
|  | Sensitivity/Specificity | 0.90 /0.70 | 0.90 /0.63 | 0.90 /0.56 |
|  |  | 0.95 /0.54 | 0.95 /0.47 | 0.95 /0.40 |
|  |  | 0.75/ 0.90 | 0.67/ 0.90 | 0.59/ 0.90 |
|  |  | 0.65/ 0.95 | 0.55/ 0.95 | 0.46/ 0.95 |
|  | Net Benefit(20/40/60) | 0.16/0.11/0.09 | 0.16/0.09/0.09 | 0.17/0.09/0.08 |

#### Geographic validation

We additionally validated our model across two different geographic regions in the US. We used 32 848 children seen in Delaware Valley sites, located in the northeastern US, as a training set and 3 343 children seen in Florida, located in the southeastern US, as a test set. We report AUROC for the geographic validation in Table 2.

#### Robustness across subpopulations

Robustness across subpopulations (group fairness) is evaluated by comparing model AUROCs in the test dataset across five groups determined by the: last WFL% category before age 2 (underweight, normal weight, overweight, obesity), race (Black, White, Asian, Other), ethnicity (Hispanic, Non-Hispanic), sex (female, male), and payer (private, public).

### 3.2 Evaluation results

We trained our deep learning model using 36 191 eligible children to predict the risk of obesity using 6 different lengths of observation windows from age 0-2 to 0-7. The observed AUROC, sensitivity, specificity, and net benefit for all prediction ages are presented in Table 2. Focusing on a popular setting that is heavily studied in the literature [11, 28, 10], we specifically present discrimination results for 0-2 years observation window and obesity prediction at age 5 in Figure 3. Notably, our model outperforms the presented baseline based on a child’s last WFL% with an AUROC and AUPRC of 0.81 and 0.61 compared with 0.71 and 0.56, respectively. Our model also dominates over other strategies (baseline, intervention for all, and intervention for none) across various net benefit threshold probabilities, with significant margins above the 15% threshold^3^ probability regime.

**Figure 3:**
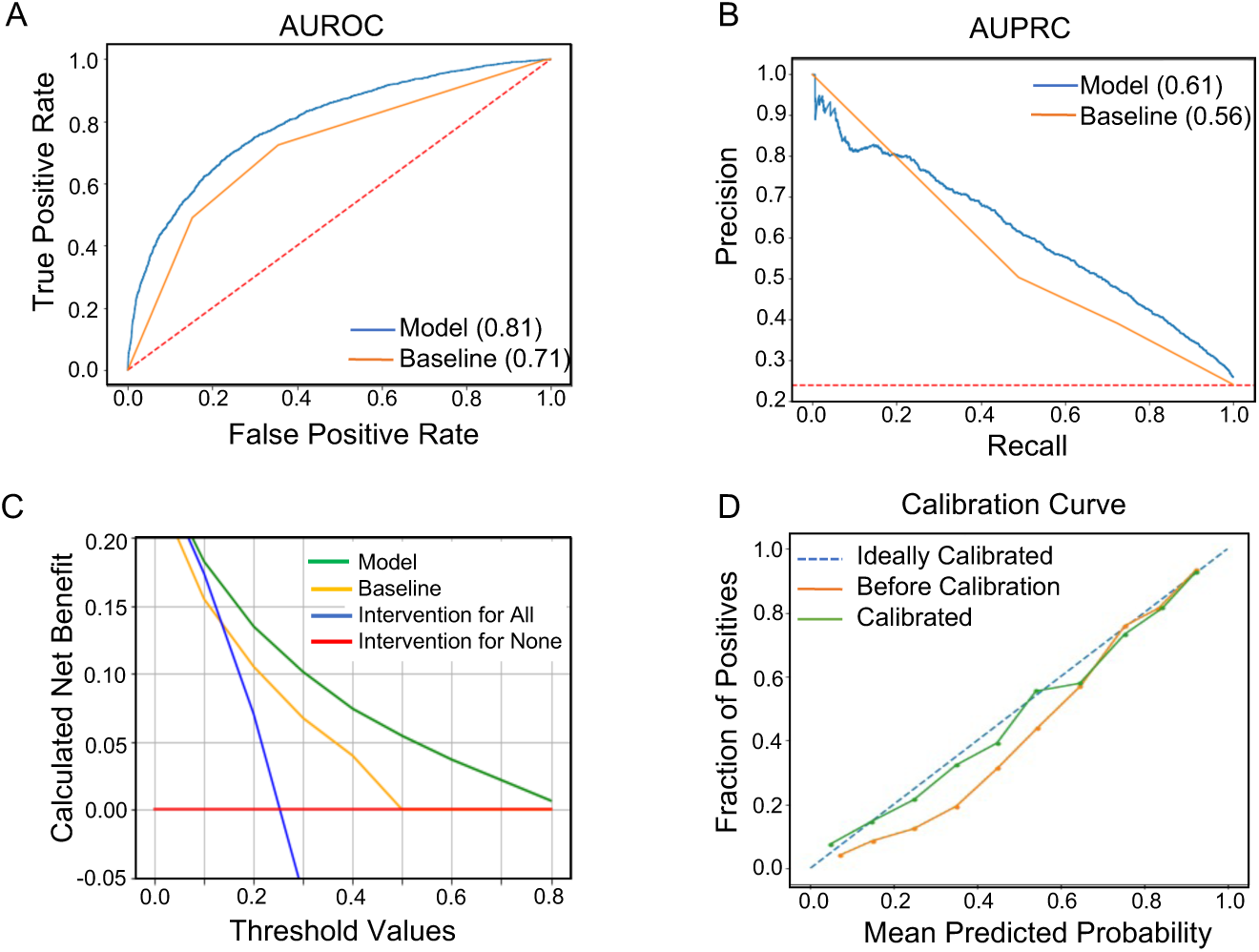
Evaluation of obesity prediction model to predict obesity at age 5 using 0-2 years of data: **A.** AUROC curve of the model (blue) and a baseline model based on the last available WFL% or BMI% measurement in the observation window (orange line), **B.** AUPRC curve of the model (blue) and a baseline model based on the last available WFL% measurement before age 2 (orange line), **C.** Decision curve analysis for different strategies of treatment, **D.** Calibration curve. The dotted line represents ideal calibration and the orange line for before calibration and the green line after calibration.

Figure 4 compares the AUROC of our model on different subpopulations of children for prediction at age 5, across the 5 groups mentioned in Section 3.1. AUROCs among each group show minimal deviation of 0.04, 0.04, 0.01, 0.001, and 0.008, respectively, showing the robustness of our model across each group.

**Figure 4:**
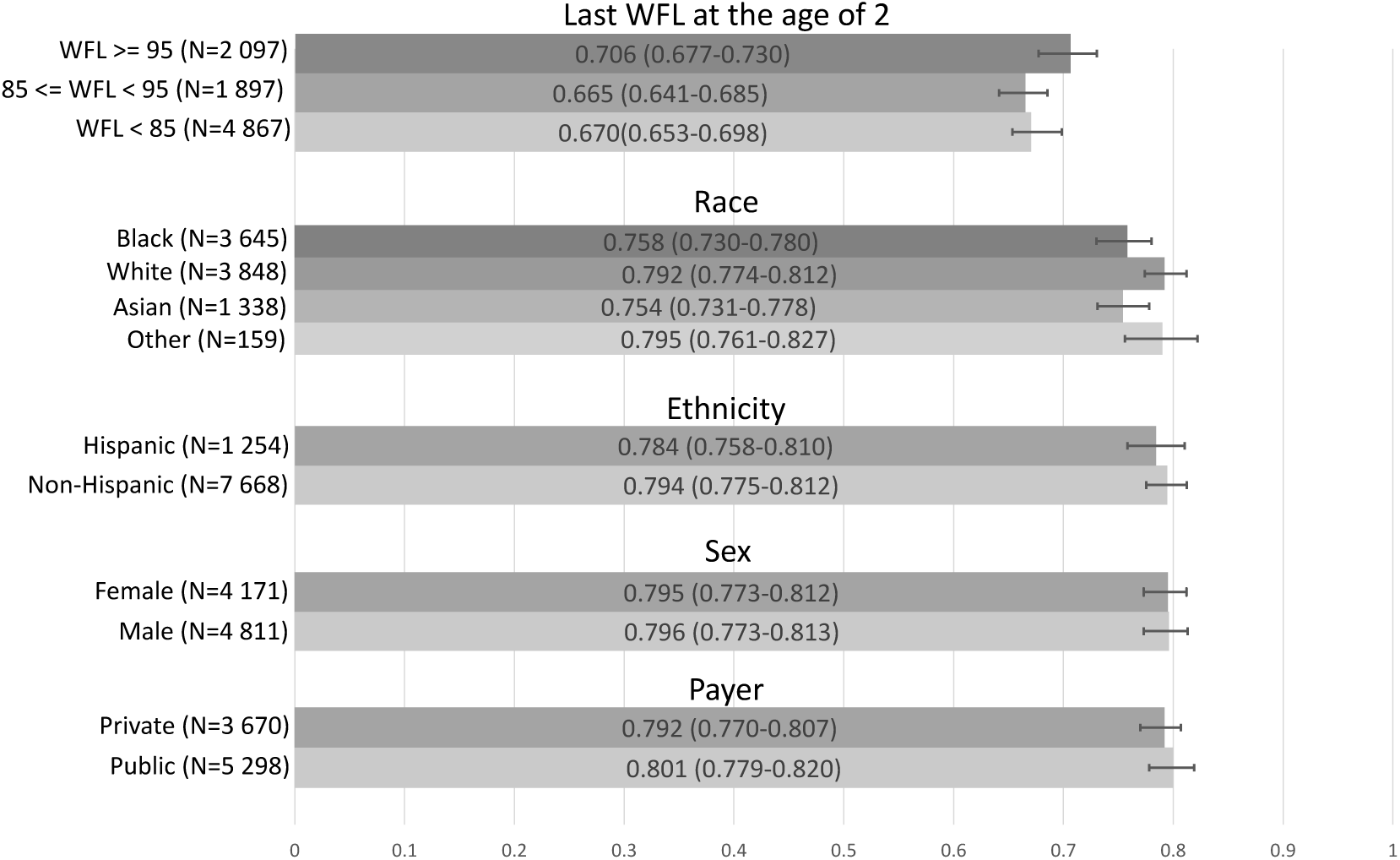
Evaluating the predictive model’s robustness by comparing AUROCs (in the test dataset) across five groups (13 subgroups): last WFL before age 2 (3 categories), race (Asian, Black, White, Other), ethnicity (Hispanic, Non-Hispanic), sex (female, male), and payer (private, public).

### 3.3 Analyzing model predictors

We investigated the risk factors identified by the model by analyzing which predictors most attribute to the model’s prediction. We used the attention scores [32, 33] obtained from the LSTM cells, as a way to determine which features are given more attention (importance) by the model to predict the output (more details are provided in Supplementary D.1).

We present the mean of the importance scores of all features inside each of the 7 input feature categories (diagnoses, family-history diagnoses, medications, measurements, demographics, last obesity status before age 2, and WFL% changes before age 2 for the entire cohort) in Figure 5.A. Clinical diagnoses and WFL% changes were the top 2 important predictor groups. We also present the ranking for the top 20 predictors based on the mean importance scores for the predictors in the entire cohort in Figure 5.B. Previous weight percentile measurements and weight gain patterns were among the top 5 features; in addition to weight-related diagnoses and diagnoses of elevated blood pressure, gastroesophageal reflux disease, developmental delay, hypothyroidism, and asthma. Among the family history diagnoses, a family history of hypertension, cardiac disorder, and depression was important. Our engineered feature (WFL% change before age 2) was also among the top 10 predictors.

**Figure 5:**
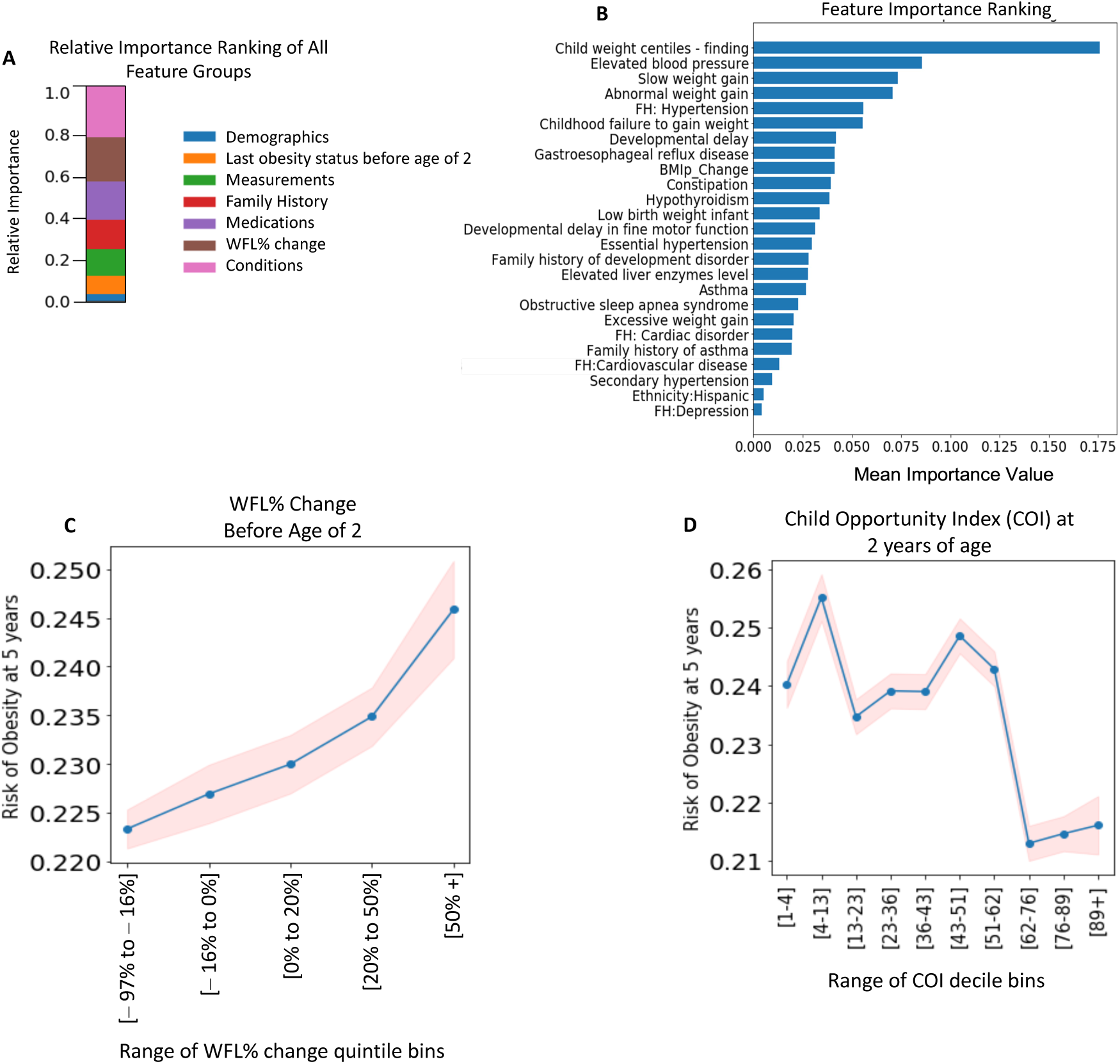
Feature Importance Analysis for all samples to predict obesity at age 5 using 0-2 years of data. **(A)** Feature importance ranking of 7 feature groups, **(B)** Relative feature importance ranking of top 20 features, **C-D** partial dependence plots between risk for obesity and **(C)** Change in WFL% before age 2 over 5 percentile categories (Dividing the numeric data (WFL%) into 5 bins (aka quintiles) such that there are an equal number of observations in each bin. This would produce a categorical object indicating quantile membership for each data point. The x-axis shows the range of each bin.), **(D)** child opportunity index (COI) at age 2 over COI-decile (Dividing the numeric data into 10 bins (aka deciles) such that there are an equal number of observations in each bin. This would produce a categorical object indicating decile membership for each data point. The x-axis shows the range of each bin.) categories.

We also evaluated partial dependency plots (PDP^4^) for the WFL% change before age 2, and child opportunity index, as a function of the predicted risk of (the probability of developing) obesity at age 5 for the test dataset. Specifically, Figure 5.C shows that an increase in the WFL% before age 2 increases the risk of obesity at age 5. Figure 5.D shows that the risk of obesity decreases with increasing COI score.

## 4. Discussion

In this study, we developed and extensively validated a model that can predict obesity reliably in early childhood using a large EHR dataset with over 36 000 children using deep learning methods. Our descriptive analysis in Section 2.3 underscores the importance of having a tool that includes other elements beyond a static WFL% or BMI% to identify children at high-risk for developing obesity before their WFL% or BMI% goes above the 95^th^ percentile. Our model used around 500 clinically relevant features from the EHR and demonstrated strong performance across multiple age ranges, chronological time periods, geographies, and demographic subgroups. Because our model is able to leverage unaugmented EHR data collected as part of routine clinical care and provides flexibility in the age at which the model is applied, there is potential for this model to be integrated into common EHRs to provide clinical decision support at multiple time points and support the implementation of preventive measures before a child develops obesity between 3 to age 10.

To date, there has been limited research on using deep learning methods with large longitudinal EHR data to predict childhood obesity [34, 12, 18]. There is a large body of studies that use logistic regression to predict the development of obesity at a certain age, most commonly 5 years. Three of the most comparable ones are presented by Redsell et al. [35], Robson et al. [36], and Hammond et al. [11] with AUROC of 0.67, 0.78, and 0.81, respectively. While these studies used a smaller set of features, they also used features like parental weight, parental smoking habits, prenatal history, and infant dietary habits which are not available in many EHRs. Several advanced ML-based models were compared by Dugan et al. [37] and Pang et al. [24] using 167 features (anthropometric measures and questionnaire data), and 102 features (maternal and EHR data), respectively for children before age 2. They reported an accuracy of 0.85 using a decision-tree method at age 3 and an AUROC of 0.81 b using XGBoost for ages 2-7.

Our study improves on prior studies in several ways. Our predictive model achieves an AUROC of 0.85 (0.84-0.86), 0.83 (0.82-0.84), and 0.81 (0.80-0.82) for predicting the risk of obesity at 3, 4, and age 5, respectively, using 0-2 years of data. Our predictive model also demonstrates high accuracy for estimating the risk of obesity for the next 3 years across a wide age range from 2 to 7 years of age (Table 2). Indeed, our model provides the flexibility of learning from as much data as available for patients before the time of screening, which is a method referred to as a “flexible window design” that our team developed in a prior paper [27]. Because of this flexibility, it provides a tool to screen children at different ages and with enough of a time window before the development of obesity for preventive interventions to be effective.

Beyond demonstrating the accuracy and flexibility of our model, we demonstrated the validity of our model across temporal, geographic shifts, multiple demographic strata and the last WFL% before age 2 and demonstrated our model’s sensitivity in predicting obesity. As a screening tool, having a model that is highly sensitive is preferred to ensure that young children at risk for developing obesity are identified and preventive measures like parenting interventions, lifestyle behavior counseling, and weight checks can be implemented in a timely manner. Notably, the sensitivity of the model does mean that there is a possibility of false positives; however, because obesity prevention measures are generally low-cost and beneficial for all children no matter their weight status, we believe this is an acceptable trade-off as long as discussion about a child’s risk for developing obesity is done in a family-centered, thoughtful, and positive way and prevent any weight-related stigma that could otherwise occur.

Another important strength of our model is that it utilizes only EHR data that is collected as part of routine clinical care. Our feature selection process can be applied to any EHR dataset that contains common data elements of basic demographics, diagnoses codes, medications, and measurements. This is advantageous compared to prior models which may require extensive data collection or data linkages that are not feasible outside of the research setting.

Similar to other studies [32, 38], our evaluation of the importance of the EHR features to our model also allows us to better understand what risk factors are important to a child’s risk of obesity, which can facilitate the provision of more personalized interventions. For example, stressing healthy lifestyles for infants whose family has a strong cardiovascular history could be an effective intervention based on our model. In addition to certain diagnoses and family history, our results demonstrated that change in WFL% in infancy is an important feature to calculate from EHR measurements to determine a child’s risk of obesity. Being able to learn from these longitudinal engineered and other EHR features is an advantage of our deep learning model.

We also found that the risk of obesity decreases with the increasing child opportunity index (COI), corroborating the importance of social determinants of health on childhood obesity [39, 40]. While COI may not be part of routine EHR data, it is easily calculated with information that is in the EHR (through ZIP codes) and our study demonstrates the importance of accounting for the influence of neighborhood environments on child health outcomes.

Our study does have some limitations. First, our study is retrospective and may be affected by changes in healthcare delivery over time. However, we were able to temporally validate the model to account for time shifts between children born between 2002-2009 and 2010-2015. Second, the dataset included information from a single healthcare system. Despite this, the dataset did include children from five different states in different geographic regions and we were able to validate our model between these geographic regions. Third, a large number of children were excluded due to inadequate or missing height and weight measurements before age 2 and this may have introduced sample bias. Finally, our dataset did not contain information on lifestyle behaviors, prenatal variables, or other sociodemographic variables, which we know are important to a child’s risk for the development of obesity. However, our focus was to use an EHR dataset that included only data collected during routine clinical care, to increase the potential that the model could be integrated into existing EHR systems to provide clinical decision support for providers in identifying children at risk for obesity to facilitate early prevention efforts across many pediatric healthcare settings. Our team is actively working on creating a CDS (clinical decision support) tool on the SMART on FHIR (Fast Healthcare Interoperability Resources) platform [41], allowing its wider adoption and integration in clinical practice and increasing its interoperability.

## 5. Data availability

Our code, containing the model with parameter (weight) values, is publicly available on GitHub at https://github.com/healthylaife/ObesityPrediction. Interested scholars can access the data by contacting Nemours Biomedical Research Informatics Center and signing a data use agreement.

## 6. Acknowledgements

Our study was supported by NIH awards, P20GM103446 and U54-GM104941.

## Abbreviations

AUPRC: Area Under Precision-Recall (PR) Curve
AUROC: Area Under the Receiver Operating Characteristic
BMI: Body Mass Index
CDC: Centers for Disease Control and Prevention
EHR: Electronic Health Record
US: United States
WFL: Weight-for-length
WHO: World Health Organization

## Footnotes

1 An encoder-decoder refers to a design, where an initial network (encoder) receives input and maps (compresses) that to a lower dimension representation. Then a second network (decoder) learns from the encoder output to generate output.

2 LSTMs are a type of (deep) neural network used for processing sequential data types.

3 Here, a threshold such as 10% means that for every 10 children evaluated for risk of obesity by the predictive model, the health service is willing to risk the cost of mistakenly prescribing 9 children [28].

4 Demonstrates the marginal effect of the different values of the feature of interest on the predicted outcome

