## supplementary material for "Reliable prediction of childhood obesity using only routinely collected EHRs is possible"

*Nemours Children’s Health, Wilmington, DE, USA*

**Daniel Eckrich**

*Nemours Children’s Health, Wilmington, DE, USA*

**H. Timothy Bunnell**

*Nemours Children’s Health, Wilmington, DE, USA*

**Rahmatollah Beheshti**

*Computer & Info. Sciences, and Epidemiology, University of Delaware, Newark, DE, USA*

### Appendix A. Nemours EHR

Nemours Children’s Health, is a large network of pediatric health in the US, primarily spanning the states of Delaware, Florida, New Jersey, and Pennsylvania. The dataset is a portion of the larger PEDSnet dataset, containing EHR data from several major US Pediatric Health Systems [1]. It contains direct clinical data from nearly all clinical and healthcare interactions. Our data is extracted from over two million distinct patients from the Nemours EHR system with patient records dating from 2002 to 2019. The analysis dataset was further screened for inconsistencies particularly those related to birthdates and measurement dates, dropping records with missing or implausible dates. The dataset was anonymized. All of the dates were skewed randomly per patient by +/- 180 days. The data access and processing steps were approved by the Nemours institutional review board. Each record in our dataset relates to one visit and captures the visit start and end time and all the condition, procedure, medication, and measurement features recorded for that visit. It also contains demographic data for each patient. The medical codes are standardized terminologies of SNOMED-CT, RxNorm, CPT, and LOINC [2] for both clinical and demographic facts.

From 68,029 children EHR data from Nemours, 65,725 children had weight and length measurements during at least 2 routine infant checkups before age 2, which are generally scheduled at ages 1, 3, 6, 9, 12, 18, and 24 months. Of them, 37,844 children had at least one weight and height measurement between 2 to age 10. We excluded 1,653 children whose year of birth could not be verified, leaving 36,191 patients for the construction of a prediction model for childhood obesity. The cohort was further divided according to date of birth for temporal validation: data of 26,786 children who were born between January 1, 2002, and December 31, 2009, were included as a training set, and data of 9,405 children

who were born between January 1, 2010, and December 31, 2015, were included in the temporal test set, respectively. For geographic validation cohort was divided according to the location of the facility visited: data of 32,848 children seen in Delaware Valley located in the northeastern US, were included as a training set, and data of 3,343 children seen at different facilities across Florida, located in the southeastern US, were included in the geographic test set, respectively.

### Appendix B. Feature Generation Methods

The original dataset consisted of 20,298 diagnoses, 6,077 medications, and 7,693 measure- ments (lab-results) features. On average, 22 diagnoses, 15 medications, and 49 measurement features were recorded per patient in the data. To identify a subset of clinically-relevant features to childhood obesity, we have used a data-driven approach coupled with input from a panel of childhood obesity experts from the PEDSnet Healthy Weight Network [3].

A total of 506 features were constructed from the dataset. We selected 71 clinical diagnoses codes that fall under broader categories of obesity-related comorbidities: cardiovascular, gastrointestinal, genetic, metabolic, neuropsychological, orthopedic, and pulmonary. We also selected 67 family history diagnoses available in our dataset recorded in the patient’s EHR. We grouped all medication codes in our dataset into 84 ATC-3 groups. Measure- ments features were first arranged in the decreasing order of their availability in our dataset and then with the help of clinical experts, we selected 51 measurements from the top 70 measurements with the help of clinical experts. All these steps taken to select the list of features were verified by clinical experts on our team. Please refer to Table S1 for the list of all features. The following sections describe the feature selection and generation mechanism for each data type.

#### B.1 Demographic Features

Gender, race, ethnicity, and payer information were extracted from EHR data. Each of these features is divided into categories: Gender - Male and Female, race - Black, White, Asian, and Other, ethnicity - Hispanic, and Non-Hispanic, and payer - Public and Private. We also included the Child Opportunity Index (COI) value for each patient by geolocating the last address of each patient before age 2 and mapping it to the neighborhood COI score. We used the Census Geocoder tool^1^ for geolocating. COI combines indicators of educational (e.g., early childhood education enrollment, high school graduation rate), health and environment (e.g., access to healthy food, health insurance coverage), and socioeconomic opportunity (e.g., employment rate, median household income) for all US neighborhoods. We divided the COI score into deciles (10 percentile bins) to generate 10 categorical features for the COI score. In total, we get 20 demographic features.

1. h[ttps://www.census.gov/programs-surveys/geograph](http://www.census.gov/programs-surveys/geography/technical-documentation/complete-technical-)y/tec[hnical-documentation/complete-technical-](http://www.census.gov/programs-surveys/geography/technical-documentation/complete-technical-) documentation/census-geocoder.html

#### B.2 Diagnoses

First, we used the MedDRA (MedDRA) hierarchy to group diagnoses. By grouping diag- noses, we reduced the number of diagnoses from a total of 20,298 to 1,457. We grouped condition codes with a patient count above 107 (mean patient count per condition code) us- ing pt level grouping in MedDRA and condition codes with a patient count below 107 using hlt level groupings in MedDRA. Out of 1,457 diagnoses, 71 clinical diagnoses and 67 family history diagnoses were included in the study based on input from clinical experts.

#### B.3 Medications

We used the ATC-3 codes to group 6,077 medications into 613 codes. Out of 613 ATC codes, 530 were excluded to include only medication codes that were present in more than 1% of the cohort population. The remaining 84 ATC codes were used as input features for model training.

#### B.4 Measurements

Anthropometrics measurements: Weight and height measurements were used for Weight- for-Length (WFL) and BMI% calculations for ages below 2 years and above respectively. Weight-for-Length (WFL) values were segmented into 5 windows corresponding to ages 0–4 months, 4–8 months, 8–12 months, 12–18 months, and 18–24 months. If multiple measurements were recorded for a patient within a window, the most latest value was used. Weight and height values were treated as MCAR and imputed with a carry forward of the most recent value over the 5 windows. All other measurement values before age 2 were transformed into categorical features by dividing each value into five equal percentile bins. We also converted the change in WFL% values over 0–4 months, 4–8 months, 8–12 months, 12–18 months, and 18–24 months to 5-centile bins to generate 5 categorical features. We also used the last WFL% status (underweight, normal, overweight, or obesity) at age 2 for model input. BMI% values after age 2 were categorized into - obesity, overweight, underweight, and normal categories which generated 4 features. BMI% categories (underweight, normal, overweight, or obesity) at every age after the of 2 were used as input to predict the risk at future age points.

Laboratory Measurements: We selected 51 other measurements from EHR data and seg- mented them into 5 windows similar to anthropometric measurements. We then converted each measurement value into 5 equal percentile bins to convert each value into 5 categories corresponding to each percentile bin. This segmentation lead to the generation of 255 measurement features per individual.

#### B.5 Feature Representation

All the height, weight, lab measurements, medications, and diagnoses after age 2 were segmented into 1-year time windows. Medications, diagnoses, and percentile-binned mea- surements are transformed into categorical values with 1 indicating the presence of that variable and 0 otherwise. These features were marked as not MCAR and the “missing” value was indicated by a dummy category.

All features generated from EHR data between 0 to age 2 were arranged chronologically based on the visit timestamps. There is no fixed order between features with the same timestamp. All data between 2 to age 10 which is binned into 1-year time intervals were represented as a binary vector with 1 if the value is observed in the 1-year time interval and 0 otherwise.

Table S 1: List of features

Feature Type	Feature Name

Measurements	Chloride [Moles/volume] in Serum or Plasma Measurements	Carbon dioxide, total [Moles/volume] in Serum or Plasma Measurements	Sodium [Moles/volume] in Serum or Plasma Measurements	Potassium [Moles/volume] in Serum or Plasma Measurements	Heart rate

Measurements	Oxygen saturation in Arterial blood by Pulse oximetry Measurements	Hematocrit [Volume Fraction] of Blood

Measurements	MCHC [Mass/volume] by Automated count Measurements	Leukocytes [#/volume] in Blood Measurements	Erythrocytes [#/volume] in Body fluid Measurements	MCH [Entitic mass] by Automated count

Measurements	Segmented neutrophils/100 leukocytes in Blood Measurements	Monocytes/100 leukocytes in Blood

Measurements	Erythrocytes [#/volume] in Blood by Automated count Measurements	Protein [Mass/volume] in Serum or Plasma Measurements	Lymphocytes/100 leukocytes in Cerebral spinal fluid Measurements	Leukocytes [#/volume] in Body fluid

Measurements	pH of Urine by Test strip

Measurements	Eosinophils/100 leukocytes in Cerebral spinal fluid Measurements	Bilirubin.total [Mass/volume] in Serum or Plasma Measurements	Albumin [Mass/volume] in Serum or Plasma Measurements	MCV [Entitic volume]

Measurements	Alkaline phosphatase [Enzymatic activity/volume] in Serum or Plasma

Measurements	Monocytes/100 leukocytes in Cerebral spinal fluid Measurements	Erythrocyte distribution width [Ratio] in Cord blood Measurements	Specific gravity of Urine by Test strip

Measurements	Hemoglobin [Mass/volume] in Blood Measurements	Lymphocytes/100 leukocytes in Blood Measurements	Calcium [Mass/volume] in Serum or Plasma

Measurements	Alanine aminotransferase [Enzymatic activity/volume] in Serum or Plasma

Measurements	Platelets [#/volume] in Blood

Measurements	Respiratory rate

Measurements	Urobilinogen [Mass/volume] in Urine Measurements	Monocytes [#/volume] in Blood

Measurements	Neutrophils [#/volume] in Blood Measurements	Lymphocytes [#/volume] in Blood

Measurements	Platelet mean volume [Entitic volume] in Blood by Automated count

Measurements	Erythrocytes [Presence] in Urine

Measurements	Body temperature

Measurements	Lead [Mass/volume] in Capillary blood Measurements	Systolic blood pressure

Measurements	Diastolic blood pressure

Measurements	Glucose [Mass/volume] in Serum or Plasma Measurements	Basophils [#/volume] in Blood Measurements	Eosinophils/100 leukocytes in Blood Measurements	Eosinophils [#/volume] in Blood

Measurements	Urea nitrogen [Mass/volume] in Serum or Plasma Measurements	Creatinine [Mass/volume] in Serum or Plasma Measurements	Glucose [Mass/volume] in Urine by Test strip Measurements	Carbon dioxide

Measurements	Total [Moles/volume] in Blood by calculation diagnoses	Obstructive sleep apnea syndrome

diagnoses	Gastroesophageal reflux disease

diagnoses	Constipation

diagnoses	Anxiety

diagnoses	Elevated blood pressure

diagnoses	Secondary hypertension

diagnoses	Hypothyroidism

diagnoses	Asthma

diagnoses	Pure hypercholesterolemia

diagnoses	Acanthosis nigricans

diagnoses	Anxiety disorder of childhood’

diagnoses	High hemoglobin A1c level

diagnoses	Precocious puberty

diagnoses	Developmental delay

diagnoses	Elevated liver enzymes level

diagnoses	Mixed hypercholesterolemia and hypertriglyceridemia

diagnoses	Prediabetes

diagnoses	Developmental delay in fine motor function’

diagnoses	Cholelithiasis with obstruction

diagnoses	Essential hypertension

diagnoses	Anxiety disorder

diagnoses	Prehypertension

diagnoses	Asthma-chronic obstructive pulmonary disease overlap syndrome diagnoses	Diabetes mellitus

diagnoses	Anxiety disorder of childhood OR adolescence

diagnoses	Asthmatic bronchitis

diagnoses	Hypothyroidism due to Hashimoto’s thyroiditis

diagnoses	Anxiety about body function or health,

diagnoses	H/O: hypertension

diagnoses	Developmental delay in receptive-expressive language

diagnoses	Chronic kidney disease stage 5 due to hypertension

diagnoses	Hypothalamic syndrome

diagnoses	Chronic kidney disease due to hypertension

diagnoses	Malignant essential hypertension

diagnoses	Malignant secondary hypertension

diagnoses	Constipation alternates with diarrhea

diagnoses	Systolic hypertension

diagnoses	Binge eating disorder

diagnoses	Hypothyroidism caused by drug

diagnoses	Extreme obesity with alveolar hypoventilation

diagnoses	Malignant hypertension

diagnoses	Diastolic hypertension

diagnoses	Hypothalamic hypothyroidism

diagnoses	Chronic kidney disease stage 3 due to hypertension’

diagnoses	Gastroesophageal reflux in child

diagnoses	Hypothalamic overactivity

diagnoses	Primary hypertriglyceridemia

diagnoses	Transient hypertension

diagnoses	Diabetes mellitus associated with pancreatic disease

diagnoses	Hypothalamic obesity,

diagnoses	Abnormal blood pressure

diagnoses	Abnormal weight gain

diagnoses	Obesity

diagnoses	Childhood failure to gain weight

diagnoses	’Low birth weight infant

diagnoses	Failure to gain weight

diagnoses	Extremely low birth weight infant

diagnoses	Very low birth weight infant

diagnoses	Unintentional weight loss

diagnoses	Excessive weight gain

diagnoses	Slow weight gain

diagnoses	Unable to weight-bear

diagnoses	Child weight centiles - finding

diagnoses	Baby birth weight 2.0-2.5kg

diagnoses	Recent weight loss

diagnoses	Abnormal weight loss

diagnoses	Disorder relating to short gestation AND/OR low birthweight diagnoses	Birth weight 1000-2499 g

diagnoses	Excessive weight loss

diagnoses	Difficulty weight-bearing

diagnoses	Body weight AND/OR growth problem

FH diagnoses	FH: Diabetes mellitus,

FH diagnoses	FH: Cardiovascular disease

FH diagnoses	Family history of asthma

FH diagnoses	Family history of genetic disorder carrier FH diagnoses	FH: Gastrointestinal disease

FH diagnoses	FH: Thrombosis

FH diagnoses	FH: Drug dependency

FH diagnoses	Family history of cardiovascular disease in first degree male rela- tive less than 55 years of age

FH diagnoses	FH: Myocardial infarction

FH diagnoses	Family history of sudden cardiac death’ FH diagnoses	Family history of kidney disease

FH diagnoses	Family history of chronic renal impairment FH diagnoses	Family history of neurological disorder

FH diagnoses	Family history of cancer

FH diagnoses	FH: Hypercholesterolemia

FH diagnoses	Family history of autism

FH diagnoses	Family history of ischemic heart disease’ FH diagnoses	Family history of mental disorder

FH diagnoses	Family history of learning disability

FH diagnoses	’FH: Depression’

FH diagnoses	Family history of victim of physical abuse FH diagnoses	Family history of problem behavior

FH diagnoses	Family history of thromboembolic disorder, FH diagnoses	Family history of development disorder

FH diagnoses	FH: Thyroid disorder

FH diagnoses	FH: Muscular dystrophy

FH diagnoses	Family history of diabetes mellitus type 1 FH diagnoses	Family history of hyperlipidemia

FH diagnoses	Family history of alcoholism

FH diagnoses	FH: Cardiac disorder

FH diagnoses	Family history of intellectual disability FH diagnoses	Family history of disorder of pancreas

FH diagnoses	Family history of blood coagulation disorder

FH diagnoses	Family history of malignant neoplasm of thyroid

FH diagnoses	Family history of attention deficit hyperactivity disorder FH diagnoses	Family history of substance abuse

FH diagnoses	Family history of stroke

FH diagnoses	Family history of Von Willebrand disease FH diagnoses	Family history of heart failure

FH diagnoses	Family history of autism in sibling

FH diagnoses	FH: Obesity

FH diagnoses	Family history of conduction disorder of the heart FH diagnoses	FH: Hypertension

FH diagnoses	Family history of consanguinity

FH diagnoses	Family history of aneurysm of thoracic aorta

FH diagnoses	FH: Neoplasm of CNS

FH diagnoses	FH: Cardiomyopathy

FH diagnoses	Family history of polycystic ovary syndrome FH diagnoses	Family history of coronary arteriosclerosis FH diagnoses	FH: Diabetes mellitus in first degree relative FH diagnoses	FH: premature coronary heart disease

FH diagnoses	FH: Hypothyroidism

FH diagnoses	Family history of neoplasm

FH diagnoses	FH: Obstetric problem

FH diagnoses	Family history of Graves disease

FH diagnoses	’FH: Manic-depressive state

FH diagnoses	Family history of Hashimoto thyroiditis FH diagnoses	FH: Raised blood lipids

FH diagnoses	’Family history of Marfan syndrome’

FH diagnoses	Family history of cardiac arrhythmia FH diagnoses	Family history of stenosis of aortic valve FH diagnoses	Family history of fragile X syndrome

FH diagnoses	Family history of diabetes mellitus type 2 FH diagnoses	Family history of smoking’

FH diagnoses	’FH: FH diagnoses anomaly

FH diagnoses	Family history of aneurysm of artery

FH diagnoses	FH: Diabetes in pregnancy

BMI% Labels	Obesity

BMI% Labels	Overweight

BMI% Labels	Underweight

BMI% Labels	Normal

Medication-ATC	Beta-lactam Antibacterials, Penicillins Medication-ATC	Other Analgesics And Antipyretics

Medication-ATC	Agents For Treatment Of Hemorrhoids And Anal Fissures For Topical Use

Medication-ATC	Adrenergics, Inhalants Medication-ATC	Antifungals For Topical Use Medication-ATC	Corticosteroids, Plain

Medication-ATC	Antiinfectives And Antiseptics, Excl. Combinations With Corti- costeroids

Medication-ATC	Antiinfectives

Medication-ATC	Corticosteroids For Systemic Use, Plain

Medication-ATC	Drugs For Peptic Ulcer And Gastro-oesophageal Reflux Disease (gord)

Medication-ATC	Other Beta-lactam Antibacterials

Medication-ATC	Antiinflammatory And Antirheumatic Products, Non-steroids Medication-ATC	Throat Preparations

Medication-ATC	Antihistamines For Systemic Use

Medication-ATC	Other Drugs For Obstructive Airway Diseases, Inhalants Medication-ATC	Antibiotics For Topical Use

Medication-ATC	Intestinal Antiinfectives

Medication-ATC	Macrolides, Lincosamides And Streptogramins Medication-ATC	Hypnotics And Sedatives

Medication-ATC	I.v. Solutions

Medication-ATC	Antivaricose Therapy

Medication-ATC	Corticosteroids And Antiinfectives In Combination Medication-ATC	Vitamin A And D, Incl. Combinations Of The Two Medication-ATC	Drugs For Constipation

Medication-ATC	Decongestants And Other Nasal Preparations For Topical Use Medication-ATC	Lipid Modifying Agents, Plain

Medication-ATC	Sulfonamides And Trimethoprim Medication-ATC	Opioids

Medication-ATC	Anxiolytics

Medication-ATC	Anesthetics, General Medication-ATC	Vitamin B12 And Folic Acid Medication-ATC	Adrenergics For Systemic Use Medication-ATC	I.v. Solution Additives Medication-ATC	Stomatological Preparations Medication-ATC	Antiemetics And Antinauseants Medication-ATC	Other Mineral Supplements

Medication-ATC	Nasal Decongestants For Systemic Use Medication-ATC	Other Nervous System Drugs Medication-ATC	Antacids

Medication-ATC	Other Systemic Drugs For Obstructive Airway Diseases Medication-ATC	Antiinflammatory/antirheumatic Agents In Combination Medication-ATC	Sodium Chloride 0.111 Meq/ml Nasal Spray [saline Spray] Medication-ATC	Anesthetics, Local

Medication-ATC	Other Otologicals

Medication-ATC	Cardiac Stimulants Excl. Cardiac Glycosides Medication-ATC	Antifungals For Systemic Use

Medication-ATC	Iron Preparations

Medication-ATC	Petrolatum Topical Ointment

Medication-ATC	Drugs For Functional Gastrointestinal Disorders Medication-ATC	Propulsives

Medication-ATC	Other Dermatological Preparations Medication-ATC	Sodium Chloride Nasal Spray Medication-ATC	Antiarrhythmics, Class I And Iii Medication-ATC	Corticosteroids, Other Combinations

Medication-ATC	Antipruritics, Incl. Antihistamines, Anesthetics, Etc. Medication-ATC	Simethicone Oral Suspension

Medication-ATC	Antidiarrheal Microorganisms Medication-ATC	Sodium Chloride Nasal Solution Medication-ATC	High-ceiling Diuretics Medication-ATC	Direct Acting Antivirals Medication-ATC	Antiepileptics

Medication-ATC	Antimycotics For Systemic Use

Medication-ATC	Muscle Relaxants, Peripherally Acting Agents Medication-ATC	Corticosteroids, Combinations With Antibiotics Medication-ATC	Decongestants And Antiallergics

Medication-ATC	Other Antibacterials

Medication-ATC	Antiglaucoma Preparations And Miotics Medication-ATC	Ectoparasiticides, Incl. Scabicides Medication-ATC	Levalbuterol Inhalation Solution Medication-ATC	Irrigating Solutions

Medication-ATC	Antispasmodics In Combination With Psycholeptics Medication-ATC	Antiinflammatory Agents And Antiinfectives In Combination Medication-ATC	Chemotherapeutics For Topical Use

Medication-ATC	Racepinephrine Inhalation Solution Medication-ATC	Immunoglobulins

Medication-ATC	Aminoglycoside Antibacterials Medication-ATC	Antithrombotic Agents

Medication-ATC	Vitamin K And Other Hemostatics Medication-ATC	Gadopentetate Dimeglumine Prefilled Syringe Medication-ATC	Other Diagnostic Agents

Medication-ATC	Antiseptics And Disinfectants Medication-ATC	Parasympathomimetics Medication-ATC	Urologicals

Medication-ATC	Dextromethorphan Hydrobromide / Pseudoephedrine Hydrochlo- ride

### Appendix C. Model Architecture

We adopted the recurrent neural network encoder-decoder architecture presented in our previous work [4]. Encoder is the neural network consisting of embedding layers (256- dimension) for diagnoses, medications, procedures, and measurements, two layers of LSTM cells (512-dimension). As shown in Figure S1, the LSTM encoder takes 0-2 years of EHR data as input and outputs its representation vector. All the features in 0-2 years of EHR data are arranged chronologically, where the order of events occurring at the same timestamps is random. The output representation vector obtained from the LSTM encoder is concatenated with demographic data representation. Demographic data is embedded into latent space using an embedding layer (256-dimension). This concatenated vector is given to the decoder as input. The decoder concatenates this vector with the EHR data from 3 to 7 years as applicable. The decoder can learn from different lengths of medical data. For example, if a patient had EHR data from 0 to 3 years of age, their EHR data vector for the third year is combined with the vector representation derived from the encoder. This combined vector is then used by the decoder network to predict the risk of obesity for the next 1, 2, and 3 years. the Decoder architecture in our previous work [4] is modified to contain three separate feed-forward networks with two fully-connected layers (512-dimension with

Output Risk of Obesity

Feed- Forward Decoder

Output Risk of Obesity

Feed- Forward Decoder

Temporal Data Representation

LSTM Encoder

0-2 years EHR data arranged chronologically

Figure S 1: Model Architecture. The dotted block is optional and is required for data after age 2.

Output Risk of Obesity

Feed- Forward Decoder

Demographic Data Representation

Embedding Layer

Demographic Data

Concatenate EHR data after 2 years

Attention Layer

Concatenated Vector

leakyRelu of 0.1, 256-dimension, 0.2 dropout) for every future age-point for the next three years. There is a third sigmoid layer at the end of two fully-connected layers to give the final output. Each of these three separate feed-forward networks is used to simultaneously provide the risk of obesity for every future age-point in the next three years.

Attention is applied to the output of encoder LSTM to rank features in it [5]. Attention layers are non-linear feed-forward layers that give softmax scores to input features in the vector representation. This will help evaluate the model’s interpretation by analyzing the ranking of softmax scores given to the features.

### Appendix D. More on experiments

#### D.1 Attentions scores

Bahdanau et al. [5] proposed this attention mechanism to automatically (soft-)search for parts of the input data that are relevant to predicting the output and assign an attention score to the predictors in the input data. Attention scores have been used in various clinical prediction models ([6, 7, 8, 9]) to provide interpretation into what predictors are more important in predicting the output of the model. A predictor’s attention score represents the weightage given to that predictor to predict the final output. These attention scores are used to obtain the weighted sum of all predictor latent representations to obtain the final representation of the patient which is then used to predict the risk of obesity. This method

enables us to capture the nonlinear relation between a predictor’s impact on the prediction. We can therefore analyze predictor attributions at the individual level, by examining the attention score of the predictors. An analysis of predictor attribution was performed using attention scores from attention layers in the LSTM model [5].

[//pedsnet.org/data/common-data-model](https://pedsnet.org/data/common-data-model).

[3] Erinn T Rhodes, Thao-Ly T Phan, Elizabeth R Earley, Ihuoma Eneli, Matthew A Haemer, Nikki C Highfield, Saba Khan, Grace Kim, Shelley Kirk, Elizabeth Monti Sullivan, et al. Patient-Reported Outcomes to Describe Global Health and Family Relationships in Pediatric Weight Management. *Childhood Obesity*, 2023.
[4] Mehak Gupta, Raphael Poulain, Thao-Ly T. Phan, H. Timothy Bunnell, and Rahmatol- lah Beheshti. Flexible-Window Predictions on Electronic Health Records. *Proceedings of the AAAI Conference on Artificial Intelligence*, 36(11):12510–12516, Jun. 2022. doi: 10.1609/aaai.v36i11.21520. URL [https://ojs.aaai.org/index.php/AAAI/article/](https://ojs.aaai.org/index.php/AAAI/article/view/21520) [view/21520](https://ojs.aaai.org/index.php/AAAI/article/view/21520).
[5] Dzmitry Bahdanau, Kyunghyun Cho, and Yoshua Bengio. Neural machine translation by jointly learning to align and translate. *arXiv preprint arXiv:1409.0473*, 2014.
[6] Junyu Luo, Muchao Ye, Cao Xiao, and Fenglong Ma. Hitanet: Hierarchical time-aware attention networks for risk prediction on electronic health records. In *Proceedings of the 26th ACM SIGKDD International Conference on Knowledge Discovery & Data Mining*, pages 647–656, 2020.
[7] Fenglong Ma, Radha Chitta, Jing Zhou, Quanzeng You, Tong Sun, and Jing Gao. Dipole: Diagnosis prediction in healthcare via attention-based bidirectional recurrent neural networks. In *Proceedings of the 23rd ACM SIGKDD international conference on knowledge discovery and data mining*, pages 1903–1911, 2017.
[8] Enliang Xu, Shiwan Zhao, Jing Mei, Eryu Xia, Yiqin Yu, and Songfang Huang. Multiple mace risk prediction using multi-task recurrent neural network with attention. In *2019 IEEE International Conference on Healthcare Informatics (ICHI)*, pages 1–2. IEEE, 2019.
[9] Jiebin Chu, Wei Dong, Kunlun He, Huilong Duan, and Zhengxing Huang. Using neu- ral attention networks to detect adverse medical events from electronic health records. *Journal of biomedical informatics*, 87:118–130, 2018.
